# Aerosol and Surface Transmission Potential of SARS-CoV-2

**DOI:** 10.1101/2020.03.23.20039446

**Authors:** Joshua L. Santarpia, Danielle N. Rivera, Vicki L. Herrera, M. Jane Morwitzer, Hannah M. Creager, George W. Santarpia, Kevin K. Crown, David M. Brett-Major, Elizabeth R. Schnaubelt, M. Jana Broadhurst, James V. Lawler, St. Patrick Reid, John J. Lowe

**Affiliations:** University of Nebraska Medical Center; National Strategic Research Institute; United Stated Air Force School of Aerospace Medicine

## Abstract

The novel severe acute respiratory syndrome coronavirus 2 (SARS-CoV-2) originated in Wuhan, China^4^ in late 2019, and its resulting coronavirus disease, COVID-19, was declared a pandemic by the World Health Organization on March 11, 2020. The rapid global spread of COVID-19 represents perhaps the most significant public health emergency in a century. As the pandemic progressed, a continued paucity of evidence on routes of SARS-CoV-2 transmission has resulted in shifting infection prevention and control guidelines between clasically-defined airborne and droplet precautions. During the initial isolation of 13 individuals with COVID-19 at the University of Nebraska Medical Center, we collected air and surface samples to examine viral shedding from isolated individuals. We detected viral contamination among all samples, indicating that SARS-CoV-2 may spread through both direct (droplet and person-to-person) as well as indirect mechanisms (contaminated objects and airborne transmission). Taken together, these finding support the use of airborne isolation precautions when caring for COVID-19 patients.

## Main Text

Healthcare worker protection and effective public health measures for emerging infectious diseases require guidance based upon a solid understanding of modes of transmission. Scant evidence describing SARS-CoV-2 transmission dynamics has led to shifting isolation guidelines from the WHO, U.S. CDC and other public health authorities. Evidence suggests that other emerging coronavirus diseases (e.g. SARS and MERS) have airborne transmission potential^5, 6^ in addition to more direct contact and droplet transmission. At least one study suggests that MERS-CoV has the possibility of transmission from mildly ill or asymptomatic individuals.^7^ Surface samples taken in patient care areas for MERS-CoV and SARS-CoV have shown positive PCR results^6^; however, experts question the presence of viable virus and the implication for transmission through fomites contaminated by the direct contact of the infected person or the settling of virus-laden particles onto the surface.^8^ Nonetheless, nosocomial outbreaks suggest transmission of coronaviruses via environmental contamination.^9,10^ While nosocomial transmission of SARS-CoV-2 is reported, the role of aerosol transmission and environmental contamination remains unclear, and infection preventionists require further data to inform appropriate practices. ^11^

The University of Nebraska Medical Center (UNMC), with its clinical partner Nebraska Medicine, cared for 13 individuals with confirmed SARS-CoV-2 infection evacuated from the Diamond Princess cruise ship as of March 6^th^, 2020. Patients requiring hospital care were managed in the Nebraska Biocontainment Unit (NBU), and mildly ill individuals were isolated in the National Quarantine Unit (NQU), both located on the medical center campus. Key features of the NBU and NQU include: 1) individual rooms with private bathrooms; 2) negative-pressure rooms (> 12 ACH) and negative-pressure hallways; 3) key-card access control; 4) unit-specific infection prevention and control (IPC) protocols including hand hygiene and changing of gloves between rooms; and 5) personal protective equipment (PPE) for staff that included contact and aerosol protection.^12^

We initiated an ongoing study of environmental contamination obtaining surface and air samples in 2 NBU hospital and 9 NQU residential isolation rooms housing individuals testing positive for SARS-CoV-2. Samples were obtained in the NQU on days 5-9 of occupancy and in the NBU on day 10. Additional samples were obtained in the NBU on day 18, after Patient 3 had been admitted to the unit for four days. We obtained surface samples, high-volume (50 Lpm) air samples, and low-volume (4 Lpm) personal air samples. The surface samples came from common room surfaces, personal items, and toilets. Personal air sampling devices were worn by study personnel on two days during sampling of NBU and NQU rooms.

During the sampling, individuals in isolation were recording symptoms and oral temperatures twice a day. The maximum temperature, during the three days preceding sampling, was recorded, as was the presence of any symptoms. During this time, 57.9% of patients recorded a temperature greater than 99.0 F, and 15.8% had a temperature greater than 100F. Independent of temperature, 57.9% of patients reported other symptoms, primarily cough.

Surface and aerosol samples were analyzed by RT-PCR targeting the E gene of SARS-CoV-2.^13^ Of the 163 samples collected in this study, 121 (72.4%) had a positive PCR result for SARS-CoV-2. Due to the need to cause minimal disruption to individuals in isolation and undergoing hospital care, the precise surface area sampled in this study was not uniform, so results are presented as concentration of gene copies present in the recovered liquid sample (copies/µL). Viral gene copy concentrations recovered from each sample type were generally low and highly variable from sample to sample ranging from 0 to 1.75 copies/µL (Figure 1A, and Tables S1 and S2), with the highest concentration recovered from an air handling grate in the NBU. Both the sampling time and flow rate were known for all aerosol samples collected in this study, therefore the airborne concentration was calculated for all air samples (copies/L of air; Table S1 and S2).

**Figure 1.**
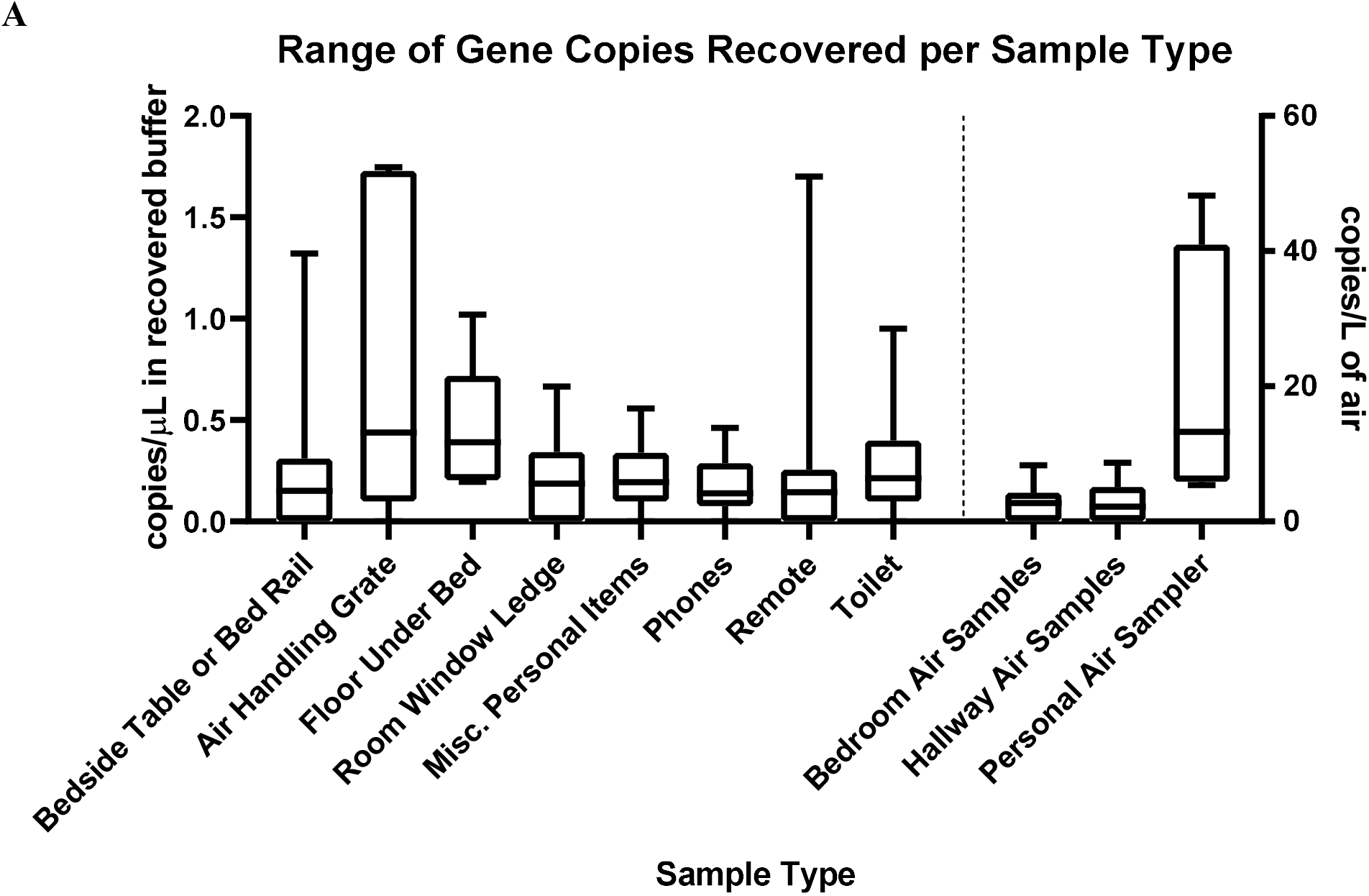
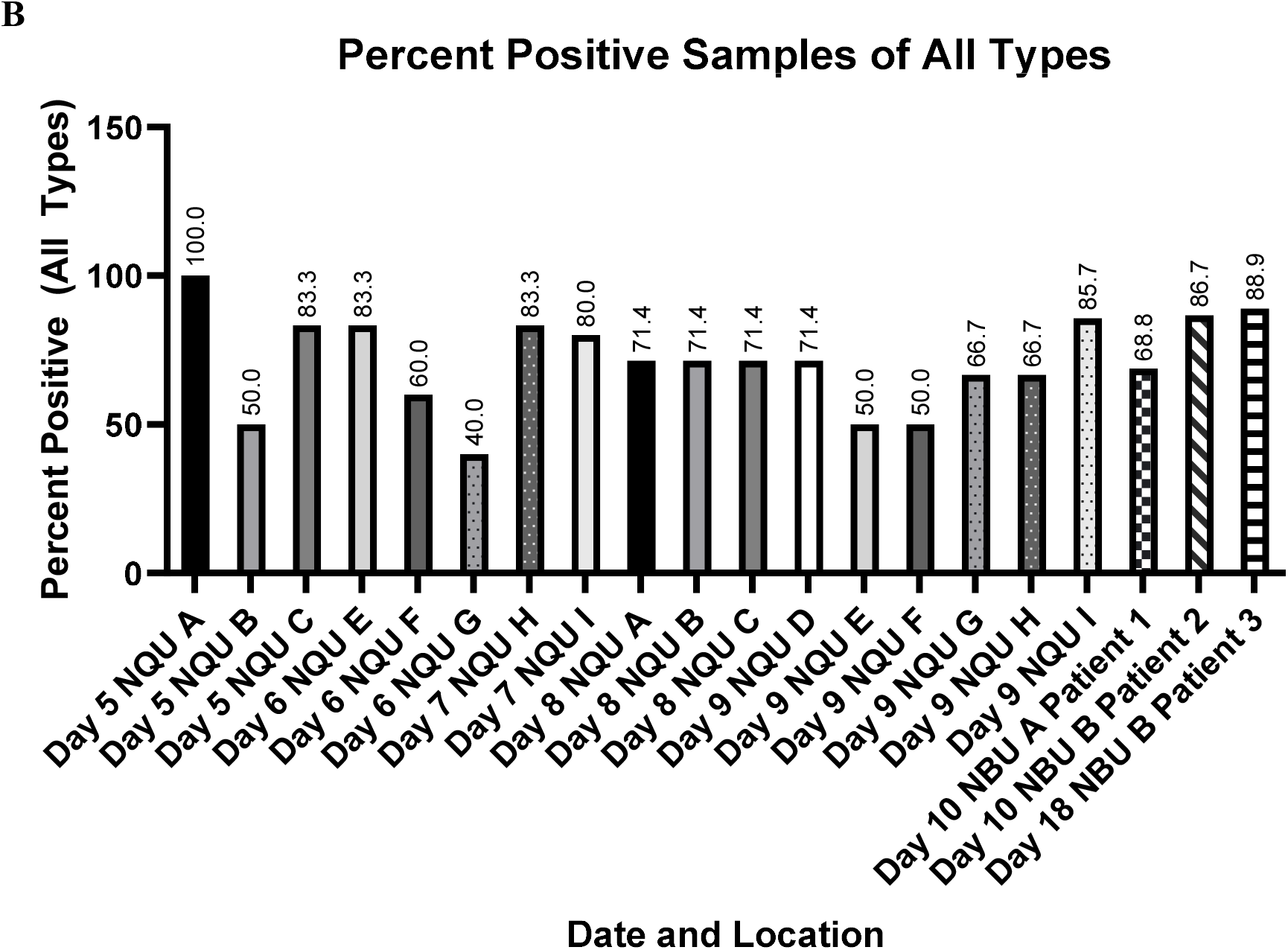
**A**. Box and whisker plot demonstrating the max and min (whiskers), median (line) and 25^th^ and 75^th^ percentile gene copy concentrations (copies/µL) for all types of samples collected in this study. Data is presented as a concentration in recovered buffer (sterile PBS) for each sample. Surface samples were in a total of 18 mL (3 mL to pre-moisten and 15 mL to recover), bedroom air and hallway air samples were recovered in 15mL total, while personal air samples were recovered in 10 mL of sterile PBS. **B**. Percentage of positive samples recovered in each room sampling. Bar patterns from the same room and same individual on multiple dates are identical.

Overall, 70.6% of all personal items sampled were determined to be positive for SARS-CoV-2 by PCR (Figure 1B and Table S1). Of these samples, 75.0% of the miscellaneous personal items (described in the methods) were positive by PCR, with a mean concentration of 0.22 copies/µL. Samples of cellular phones were 77.8% positive for viral RNA (0.17 copies/µL mean concentration) and remote controls for in-room televisions were 55.6% percent positive (mean of 0.22 copies/µL). Samples of the toilets in the room were 81.0% positive, with a mean concentration of 0.25 copies/µL. Of all room surfaces sampled (Figure 1B and Table S1), 75.0% were positive for SARS-CoV-2 RNA. 70.8% of the bedside tables and bed rails indicated the presence of viral RNA (mean concentration 0.26 copies/µL), as did 72.7% of the window ledges (mean concentration 0.22 copies/µL) sampled in each room. The floor beneath patients’ beds and the ventilation grates in the NBU were also sampled. All five floor samples, as well as 4 of the 5 ventilation grate samples tested positive by RT-PCR, with mean concentrations of 0.45 and 0.82 copies/µL, respectively.

Air samples in the rooms and in the hallway spaces (Figure 1B, and Tables S1 and S2) provide information about airborne viral shedding in these facilities. We found 63.2% of in-room air samples to be positive by RT-PCR (mean concentration 2.42 copies/L of air). In the NBU, for the first two sampling events performed on Day 10, the sampler was placed on the window ledge away from the patient (NBU Room A occupied by Patient 1), and was positive for viral RNA (Table S1; 2.42 copies/L of air). During the sampling event on Day 18 in NBU Room B occupied by Patient 3, one sampler was placed near the patient and one was placed near the door greater than 2 meters from the patient’s bed while the patient was receiving oxygen (1L) via nasal cannula. Both samples were positive by PCR, with the one closest to the patient indicating a higher airborne concentration of RNA (4.07 as compared to 2.48 copies/L of air). Samples taken outside the rooms in the hallways were 58.3% positive (Figure 1B and Table S2), with a mean concentration of 2.51 copies/L of air. Both personal air samplers from sampling personnel in the NQU showed positive PCR results after 122 minutes of sampling activity (Table S2), and both air samplers from NBU sampling indicated the presence of viral RNA after only 20 minutes of sampling activity (Table S2). The highest airborne concentrations were recorded by personal samplers in NBU while a patient was receiving oxygen through a nasal cannula (19.17 and 48.22 copies/L). Neither individuals in the NQU or patients in the NBU were observed to cough while sampling personnel were in the room wearing samplers during these events.

Between 5 and 16 samples were collected from each room, with a mean of 7.35 samples per room and a mode of 6 samples per room. The percentage of positive samples from each room ranged between 40% and 100% (Figure 1B and 2A). A Spearman’s Rank Order Correlation (ρ) of percent positive samples and total number of samples for each room had a value of 0.33, indicating a weak relationship between the number of samples taken and the percent of positive samples observed, which is likely due to the focus of unplanned samples on areas or objects frequently used by patients. When the percent of positive samples taken was compared to the maximum reported oral temperature of the patient for the previous three days, a ρ of 0.39 indicated only a weak relationship between elevated body temperature and shedding of virus in the environment. Further, recorded oral temperature was compared with the gene copy concentration for each in-room sample type (Table S1). These ρ values ranged from −0.02 (cell phones) to 0.36 (air samples), indicating weak to no significant relationship between body temperature and environmental contamination, with air samples having the strongest correlation, followed by windowsills (ρ = 0.25) and remotes (ρ = 0.23).

A subset of samples that were positive for viral RNA by RT-PCR was examined for viral propagation in Vero E6 cells. Several indicators were utilized to determine viral replication including cytopathic effect (CPE), immunofluorescent staining, time course PCR of cell culture supernatant, and electron microscopy. Due to the low concentrations recovered in these samples cultivation of virus was not confirmed in these experiments. Nevertheless, in two of the samples, cell culture indicated some evidence for the presence of replication competent virus (Figure 2): an air sample from the NQU hallway on day 8 and the windowsill from NQU A on day 5 (Table S1 and S2). Microscopic inspection of cell cultures indicated CPE after 3-4 days (Figure 2A-B). Serial PCR of cell culture supernatant was unlcear, but the observed changes in supernatant RNA, in the hallway sample, indicated that after an initial decrease in RNA in the supernatant (consistent with the with daily withdrawal of supernatant for analysis and replacement with fresh supernatant) some increase in viral RNA may have occurred (Figure 2C). The windowsill sample had consistent viral RNA in the supernatant throughout the time course, despite the daily withdrawal of supernatant for analysis, which could indicate replication (Figure 2D). Further, immunofluorescence images (Figure 2E) indicate evidence of viral proteins in the hallway sample and transmission electron microscopy (TEM) of the windowsill sample (Figure 2F) confirmed the presence of intact SARS-CoV-2 virions after 3 days of cell culture.

**Figure 2.**
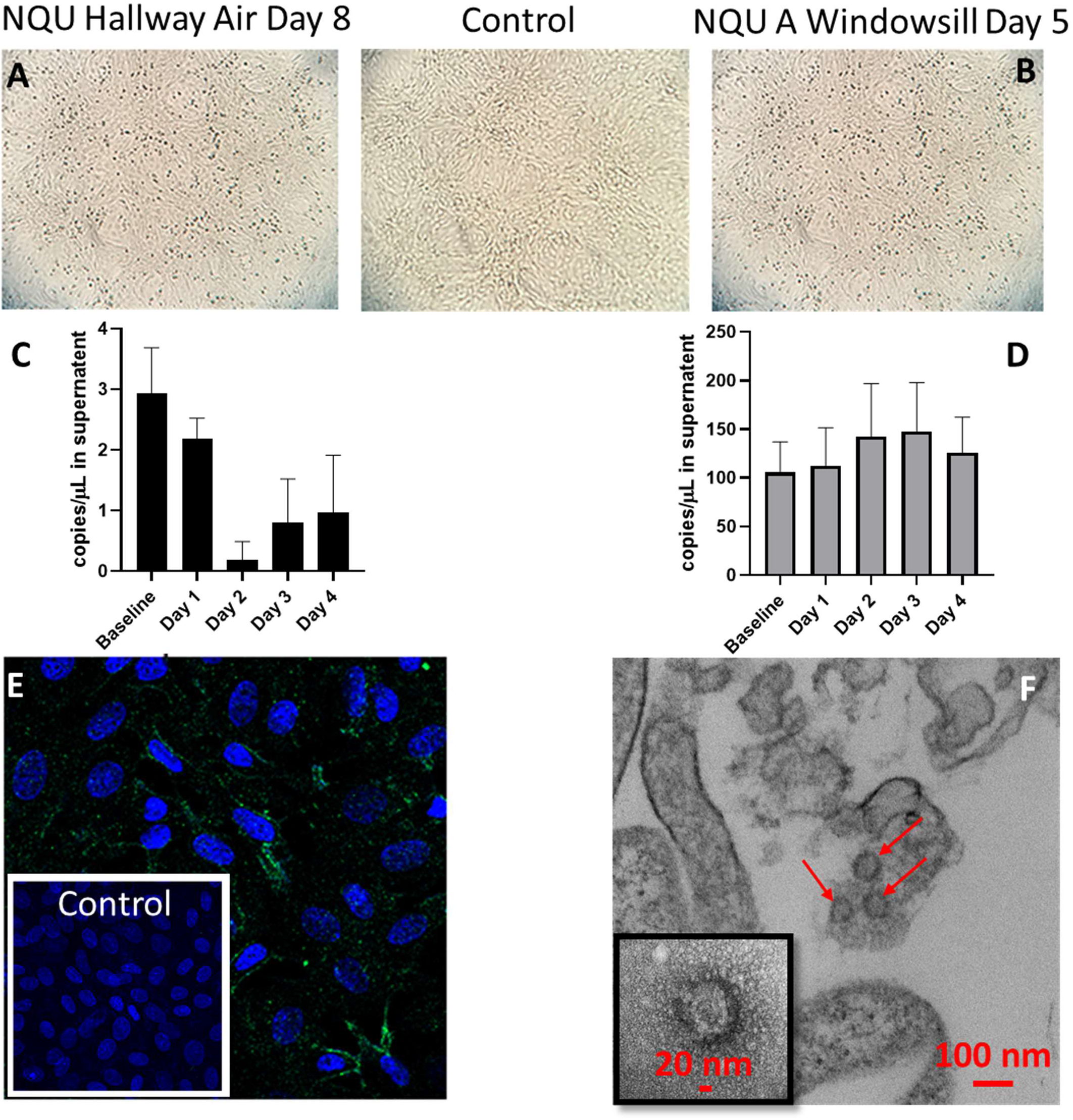
**Results of SARS-CoV-2 cell culture experiments**. Images and graph describe the results of cell culture of two environmental samples. The two samples are shown: an air sample from the NQU hallway on day 8 (A,C, and E), the windowsill from NQU A on day 1 (B,D, and F). Cytopathic effect observed in these samples (A-B) is generally mild, compared to the control (top center) which had no environmental sample added. RT-qPCR from daily withdrawals of 100 µL of supernatant from the cell culture of each sample indicates changes in viral RNA in the supernatant throughout cultivation. The hallway air sample indicates a decrease in RNA concentration in the supernatant over the first 2 days, consistent with the withdrawal of supernatant for analysis. Increase in concentration is observed on both days 3 and 4 (C). The windowsill sample showed stable and possible increasing viral concentrations for the first 3 days, despite the withdrawal of supernatant for analysis (D). Immunofluorescent staining of the hallway air sample indicates the presence of SARS-CoV-2, after 3 days of cell culture (E), as compared to control cells (inset), which were not exposed to any environmental sample. TEM images of the lysates from the windowsill culture (F) clearly indicate the presence of intact SARS-CoV-2 virions, after 3 days of cell culture.

Taken together, these data indicate significant environmental contamination in rooms where patients infected with SARS-CoV-2 are housed and cared for, regardless of the degree of symptoms or acuity of illness. Contamination exists in all types of samples: high and low-volume air samples, as well as surface samples including personal items, room surfaces, and toilets. Samples of patient toilets that tested positive for viral RNA are consistent with other reports of viral shedding in stool (14). The presence of contamination on personal items is also expected, particularly those items that are routinely handled by individuals in isolation, such as cell phones and remote controls, as well as medical equipment that is in near constant contact with the patient. The observation of viral replication in cell culture for some of the samples confirms the potential infectious nature of the recovered virus.

We noted variability in the degree of environmental contamination (as measured by the percentage of positive samples) from room to room and day to day. In general, percent positive samples in the NQU were higher on Days 5-7 (72.5%) vs Days 8–9 (64.9%). While most NQU rooms had higher percentages of positive specimens earlier in the course of illness, three of the nine rooms (NQU B,G, and I) actually had higher percentages on later days (Days 8 and 9 respectively). On average, a higher percentage of positive samples (81.4% over 3 rooms) was detected in the NBU later in the course of illness (sampled on Days 10 and 18), suggesting that patients with higher acuity of illness or levels of care may be associated with increased levels of environmental contamination. However, the lack of a strong relationship between environmental contamination and body temperature reaffirms the fact that shedding of viral RNA is not necessarily linked to clinical signs of illness.

In the hospital NBU, where patients were generally less mobile, distribution of positive samples suggests a strong influence of airflow. Personal and high-touch items were not universally positive, yet we detected viral RNA in 100% of samples from the floor under the bed and all but one window ledge (which were not used by the patient) in the NBU. Airflow in NBU suites originates from a register at the top center of the room and exits from grates near the head of the patient’s bed on either side of the room. Airflow modelling^15^ has suggested that some fraction of the airflow is directed under the patient’s bed, which may cause the observed contamination under the bed, while the dominant airflow likely carries particles away from the patient’s bed towards the edges of the room, likely passing by the windows resulting in some deposition there.

Although this study did not employ any size-fractionation techniques in order to determine the size range of SARS-CoV-2 droplets and particles, the data is suggestive that viral aerosol particles are produced by individuals that have the COVID-19 disease, even in the absence of cough. First, in the few instances where the distance between individuals in isolation and air sampling could be confidently maintained at greater than 6 ft, 2 of the 3 air samples were positive for viral RNA. Second, 58.3% of hallway air samples indicate that virus-containing particles were being transported from the rooms to the hallway during sampling activities. It is likely that the positive air samples in the hallway were caused by viral aerosol particles transported or resuspended by personnel exiting the room.^16,^ Finally, personal air samplers worn by sampling personnel were all positive for SARS-CoV-2, despite the absence of cough by most patients while sampling personnel were present. Recent literature investigating human-expired aerosol suggests that a large fraction is less than 10 µm in diameter across all types of activity (e.g. breathing, talking, and coughing^18^) and that upper respiratory illness increases production of aerosol particles (less than 10 µm)^19^. A recent study of SARS-CoV-2 stability indicates that infectious aerosol may persist for several hours and on surfaces for as long as 2 days.^20^

Our study suggests that SARS-CoV-2 environmental contamination around COVID-19 patients is extensive, and hospital IPC procedures should account for the risk of fomite, and potentially airborne, transmission of the virus. Despite wide-spread environmental contamination and limited SARS-CoV-2 aerosol contamination associated with hospitalized and mildly ill individuals, the implementation of a standard suite of infection prevention and control procedures prevented any documented cases of COVID-19 in healthcare workers, who self-monitored for 14 days after last contact with either ward and underwent two nasal swab PCR assays 24 hours apart if they reported fever or any respiratory infection symptoms. The standard IPC protocols for both the NBU and NQU includes negative pressure rooms with 12-15 air-exchanges per hour, negative pressure hallways in the suite compared to outside, strict access control, highly trained staff with well-developed protocols, frequent environmental cleaning, and aerosol-protective personal protective equipment that consisted of N95 filtering facepiece respirators in the NQU and powered air purifying respirators for patient care within the NBU. Further study is necessary to fully quantify risk.

## Data Availability

All data is included in manuscript materials

## Methods

High-touch personal items sampled included cellular phones, exercise equipment, television remotes, and medical equipment. Room surfaces tested included ventilation grates, tabletops, and window ledges. Toilet samples were obtained from the rim of the bowl. Air samples were collected both in isolation rooms and in the hallways of the NBU and NQU during sampling activities, while patients were present. Personal air samplers were worn by sampling personnel on two occasions during sampling activities: once during sampling at the NQU when 6 individual rooms were sampled, and once in the hospital NBU when one room was sampled.

Surface and personal items were collected using 3×3 sterile gauze pads 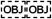pre-wetted with 3 mL of phosphate buffered saline (PBS). Large area surface samples were collected by wiping in an “S” pattern in 2 directions to cover as much of the available surface as possible. Smaller items (e.g. cellular phones, remote controls) were wiped in one direction on every available surface. Following collection, samples were packed in 50 mL conical tubes. Hand hygiene and glove changes were performed between the collection of every sample.

### Personal Items

Several personal items were sampled consistently between all quarantine rooms (cellular phones and television remote controls). In addition, individuals were asked about which items they used or handled frequently, and several additional samples were collected based on those responses: exercise equipment, medical equipment (spirometer, pulse oximeter, nasal cannula), personal computers, iPads, reading glasses, and pots used to heat water. This last category was grouped together as “Miscellaneous Personal Samples”.

### Room Surfaces

Several surfaces were sampled in each room. For rooms in the National Quarantine Unit, both the windowsill and the bedside table were sampled. For rooms in the Nebraska Biocontainment Unit, samples were taken on the windowsill, the bed rail or bedside table, under the patient’s bed and on the air conditioning return grate nearest the door.

### Air Samples

Stationary air samples, both inside and outside of patient rooms, were collected using a Sartorius Airport MD8 air sampler operating at 50 Lpm for 15 minutes. Samples were collected onto a 80mm gelatin filter. Samplers in patient rooms were placed on bedside tables and nightstands but at least 1 meter away from the patient. No attempt was made to ensure the sampler was placed a specific distance from the individual in the room, so, while distance between sampler and individual was neither defined nor consistent, individuals in the room did not directly interact with the sampler.

NQU subjects were ambulatory however, and all were out of bed during sampling. Our NQU protocol advises patients in isolation to maintain a 6-foot distance from staff members who enter their room and wear a procedure mask (e.g. surgical mask) while staff are in the room. Observed adherence with these procedures was high throughout patient stays. For this study, individuals were instructed that they could remove the mask during air sampling activities; however, many individuals did not remove it and therefore the impact of infected individuals wearing masks cannot be assessed in this study.

Study personnel generally left rooms during a significant period of time during air sampling, but it appeared that not all patients removed procedure masks during those periods. Hallway air samples were obtained by placing samplers on the floor approximately 10 cm from the door frame adjacent to rooms where sampling activities were taking place. Study personnel entered and exited rooms several times during air sampling. Additional personal air samples were collected by study personnel during sampling activities wearing Personal Button Samplers (SKC, Inc.) and using Air Chek pumps (SKC, Inc.) both sampling at 4 Lpm. These samples were collected onto 25 mm gelatin filters.

### Sample Recovery, RNA Extraction and Reverse Transcriptase PCR

Surface samples were recovered by adding 15 mL of sterile PBS to the conical vial containing the gauze pad and manually shaking the conical for 1 minute. 25 mm gelatin filters were removed from the filter housing and placed in a 50 mL conical tube and then dissolved by adding 10 mL of sterile PBS. 80 mm gelatin filters were removed from their filter housing, carefully folded and placed in a 50 mL conical tube and then dissolved by adding 15 mL of sterile PBS. RNA extractions were performed using a Qiagen DSP Virus Spin Kit (QIAGEN GMbH, Hilden, Germany) 200 to 400ul of initial sample was used for RNA extraction, and a negative extraction control was included with each set of extractions. Samples were eluted in 50ul of Qiagen Buffer AVE. RT-PCR was performed using Invitrogen Superscript III Platinum One-Step Quantitative RT-PCR System. Each PCR run included a positive synthetic DNA control and a negative, no template, control of nuclease free water. In addition, blank samples of swipes and gelatin filters, both carried during sampling, and those kept in the laboratory were analyzed. No amplification of blank samples was observed. Reactions were set up and run with initial conditions of 10 minutes at 55°C and 4 minutes at 94°C then 45 cycles of 94°C for 15 sec and 58°C for 30 seconds, QuantStudio™ 3 (Applied Biosytems™, Inc) utilizing the following reagents:

6.1 µL nuclease free water

12.5 µL Invitrogen 2X Master Mix

0.4 µl MgSO4

0.5 µl Primer/Probe Mix (IDT)* *(Primers 10uM, Probe 5uM)*

0.5 µl SuperScript III Platinum Taq

5.0 µL extracted sample RNA, nuclease free water or positive control

25.0 µL Total

In order to quantify the number of viral gene copies present in each sample from the measured Ct values, a standard curve was developed using synthetic DNA. A 6-log standard curve was run in duplicate beginning at a concentration of 1×10^3^ copies/µL. The data was fit with the exponential function:

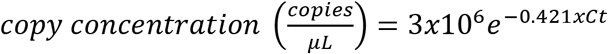

Where Ct is the cycle time where amplification is definitive. The equation was then used to convert all measured Ct values from all samples into gene copy concentrations. The minimum concentration detected by this assay was 1e^-1^ copies/µL at between 39 and 44 cycles. Considering a 5 µL sample volume and the derived exponential function above, the Ct value equating to 1 copy per reaction is 39.2, therefore, amplification beyond 39.2 is treated as undetected. The average and standard deviation concentrations were calculated from the triplicate PCR runs for each sample.

The primers and probe used in this study (below) are based on a previously published assay^13^ targeting the E gene of SARS-CoV-2, which produces the envelope small membrane protein. The gene was used as a target based on its similarity to previously identified coronavirus, including SARS-CoV strain Frankfurt and two Bat SARS-related CoV (GenBank Acc. No. MG772933.1 and NC_014470). The Primer-BLAST^21^ tool was used to examine the specificity of the assay beyond what was described in the original publication. The search used default parameters and allowed 9 mismatches before ignoring the target sequence. The search returned hits from 957 SARS-CoV-2 isolate sequences and 7 pangolin coronavirus isolate sequences, indicating that it should be sensitive SARS-CoV-2 and only have the potential to cross-react on related non-human coronaviruses. The positive control consisted of ssDNA, targeting the E and N gene (below), in a 1:1 mixture at 10^3^ copies/µL. ssDNA was based on the 2019-nCoV genome sequence published in Genbank^22^.

*E gene target primers and probe:

Probe: 5’/56-FAM/ACACTAAGCC/ZEN/ATCCTTACTGCGCTTCG/3AIBkFG/-3’

Primer 1: 5’-ATATTGCAGCAGTACGCACACA-3’

Primer 2: 5’-ACAGGTACGTTAATAGTTAATAGCGT-3’

ssDNA E Target Sequence:

5’TTCGGAAGAGACAGGTACGTTAATAGTTAATAGCGTACTTCTTTTTCTTGCTTTCGTGGTATTCTTGCTAGTTACACTAGCCATCCTTACTGCGCTTCGATTGTGTGCGTACTGCTGCAATATTGTTAACGTG-3’

ssDNA N Target Sequence:

5’ACCAAAAGATCACATTGGCACCCGCAATCCTGCTAACAATGCTGCAATCGTGCTACAACTTCCTCAAGGAACAACATTGCCAAAAGGCTTCTACGCAGAAGGGAGCAGAGGCGGCAGTCAAGCCTCTTCTCGTTCCTCATCACGTAGT-3’

## Cell Culture Assays

Vero E6 cells were used to culture virus from environmental samples. The cells were cultured in Dulbeccos’s minimal essential medium (DMEM) supplemented with heat inactivated fetal bovine 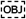serum (10%), Penicillin/Streptomycin (10,000 IU/mL &10,000 µg/mL) and Amphotericin B (25 µg/mL). For propagation, 100 µl of undiluted samples were added to 24-well plates. The cells were monitored daily to detect virus-induced CPE. After 3-4 days cell supernatants and lysates were collected. For the time course PCR experiments, supernatant was collected on each day. Samples were evaluated for cytopathic effect. Immunofluorescence, performed using mouse monoclonal SARS-CoV 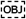to determine the presence of viral antigens. The reagent was obtained through BEI Resources, NIAID, NIH: Monoclonal Anti-SARS-CoV S Protein (Similar to 540C), NR-618. Cell nuclei were labeled with Hoechst 33342. Confocal images were collected with Ziess LSM 800 with Airyscan. For electron microscopy, samples were fixed, processed then sectioned and subsequently subjected to transmission electron microscopy (TEM). CPE images were acquired in the Nebraska Public Health Laboratory (NPHL) BSL3 facility using a 3D printed ocular adapter for cell phone photography, kindly donated by Dr. Jesse Cox, Yellow Basement Designs.

## Acknowledgments

The authors would also like to thank all of the individuals in isolation and care at both the National Quarantine Unit and the Nebraska Biocontainment Unit for their willingness and interest in cooperating with this study. The authors would like to thank Tom Bargar and Nicholas Conoan of the Electron Microscopy Core Facility (EMCF) at the University of Nebraska Medical Center for technical assistance. The EMCF is supported by state funds from the Nebraska Research Initiative (NRI) and the University of Nebraska Foundation, and institutionally by the Office of the Vice Chancellor for Research. The authors would like to thank Janice A. Taylor and James Talaska of the Advanced Microscopy Core Facility at the University of Nebraska Medical Center for providing assistance with confocal microscopy

## Funding

Funded by internal funds from the University of Nebraska Medical Center.

## Author contributions

J.S and J.J.L. conceived of the initial study; J.S. and D.R. developed the sampling strategy; J.V.L., E.S. and D.B-M. collected medical data; S.P.R. and M.J.M performed cell culture assays; G.S., J.B. and H.C. developed PCR assay and performed initial tests; J.S., J.J.L. D.R., V.H. J.V.L. collected samples; K.K.C, D.R. and V.H processed samples V.H performed all PCR; J.S. and J.J.L. wrote the manuscript with contributions from all authors.

## Declarations

This study was conducted in the National Quarantine Unit and the Nebraska Biocontainment Unit with permission from the University of Nebraska Medical Center and Nebraska Medicine as a part of a quality assurance/quality improvement study on isolation care. Sampling of individual personal items was done with the owner’s permission. Patients were informed that use of a face covering was not necessary during sampling activities, but the decision to wear or not wear provided surgical masks was left to the individual. This activity was reviewed by Office of Regulatory Affairs at the University of Nebraska Medical Center and it was determined that this project does not constitute human subject research as defined by 45CFR46.102.

## Competing interests

The authors declare no competing interests;

## Data and materials availability

All data is available in the main text or the supplementary materials

## Notes

### Competing Interest Statement

The authors have declared no competing interest.

### Funding Statement

Work was supported by University of Nebraska internal funding

## Main References

1. WHO issues a global alert about cases of atypical pneumonia. Indian J Med Sci. 2003;57(5):206–7.

2. van der Hoek L., et al. Identification of a new human coronavirus. Nat Med. 2004;10(4):368–73.

3. Woo P.C. et al. Characterization and complete genome sequence of a novel coronavirus, coronavirus HKU1, from patients with pneumonia. J Virol. 2005;79(2):884–95.

4. Zhu, N., et al. “A novel Coronavirus from patients with pneumonia in China, 2019.” New Engl J Med (2020).

5. Tellier et al. “Recognition of aerosol transmission of infectious agents: a commentary.” BMC Infect Dis. (2019) 19:101; https://doi.org/10.1186/s12879-019-3707-y

6. Booth T.F.,, et al. Detection of airborne severe acute respiratory syndrome (SARS) coronavirus and environmental contamination in SARS outbreak units. J Infect Dis. 2005; 191: 1472-1477.

7. Omrani A.S., et al. A family cluster of Middle East respiratory syndrome coronavirus infections related to a likely unrecognized asymptomatic or mild case. Int J Infect Dis. 2013;17:e668–72.

8. Morawska L. Droplet fate in indoor environments, or can we prevent the spread of infection. Indoor Air. 2006. 16(5):pp. 335–347.

9. Chowell G., et al. Transmission characteristics of MERS and SARS in the healthcare setting: a comparative study. BMC Med. 2015;13:210. doi: 10.1186/s12916-015-0450-0

10. Bin S.Y., et al. Environmental contamination and viral shedding in MERS patients during MERS-CoV outbreak in South Korea. Clin Infect Dis. 2016;62(6):755–760. doi:10.1093/cid/civ1020

11. Wang D., et al. Clinical characteristics of 138 hospitalized patients with 2019 novel coronavirus-infected pneumonia in Wuhan, China. JAMA. Published online February 7, 2020. doi:10.1001/jama.2020.1585

12. Beam, E.L., et al. Personal protective equipment processes and rationale for the Nebraska Biocontainment Unit during the 2014 activations for Ebola virus disease Am J Infect Control. Volume 44, Issue 3, 340—342

13. World Health Organization (2020, January 13). Diagnostic detection of Wuhan coronavirus 2019 by real-time RT-PCR. https://www.who.int/docs/default-source/coronaviruse/wuhan-virus-assay-v1991527e5122341d99287a1b17c111902.pdf?sfvrsn=d381fc88_2

14. Ong S.W.X., Tan Y.K., Chia P.Y. Air, Surface Environmental, and Personal Protective Equipment Contamination by Severe Acute Respiratory Syndrome Coronavirus 2 (SARS-CoV-2) From a Symptomatic Patient. JAMA. Published online March 4, 2020. doi:10.1001/jama.2020.3227

15. Hewlett A.L., et al. Mathematical modeling of pathogen trajectory in a patient care environment. Infect Control Hosp Epidemiol. 2013;34(11): 1181-1188

16. Batterman S.A. Characterization of particulate emissions from occupant activities in offices. Indoor Air., 11 (1) (2001), pp. 35–48

17. Wang, J., Chow, T.T. Numerical investigation of influence of human walking on dispersion and deposition of expiratory droplets in airborne infection isolation room. Build Environ. 2011 Oct 1;46(10):1993–2002.

18. Johnson G.R., et al. Modality of human expired aerosol size distributions. JAerosol Sci. 2011; 42: 839–851. doi:10.1016/jjaerosci.2011.07.009

19. Lee J., et al. Quantity, size distribution, and characteristics of cough generated aerosol produced by patients with an upper respiratory tract infection. Aerosol Air Qual Res. 2019; 19: 840-853.

20. van Doremalen N., et al. Aerosol and surface stability of HCoV-19 (SARS-CoV-2) compared to SARS-CoV-1. New Engl J Med. Published online March 17, 2020. DOI: 10.1056/NEJMc2004973

## Methods References

21. Ye J, Coulouris G, Zaretskaya I, Cutcutache I, Rozen S, Madden T (2012). Primer-BLAST: A tool to design target-specific primers for polymerase chain reaction. BMC Bioinformatics. 13:134.

22. GenBank. Wuhan seafood market pneumonia virus isolate Wuhan-Hu-1, complete genome. https://www.ncbi.nlm.nih.gov/nuccore/MN908947

